# Chikungunya virus (CHIKV) seroprevalence in the local population of the Cook Islands and Vanuatu and associated environmental and social factors A first CHIKV seroprevalence study in two South Pacific communities

**DOI:** 10.1101/2022.07.07.22277208

**Authors:** Charlotte E.B. Saretzki, Gerhard Dobler, Elisabeth Iro, Yin May, Douglas Tou, Eteta Lockington, Michael Ala, Nicole Heussen, Bruno S.J. Phiri, Thomas Küpper

**Affiliations:** Institute for Occupational and Social Medicine, RWTH Aachen Technical University, Aachen/ Germany. Pauwelsstraße 30, 52074 Aachen, Germany; Bundeswehr Institute of Microbiology, Munich/Germany. Neuherbergstraße 11, Ernst-von-Bergmann-Kaserne, 80937 München, Germany; Cook Islands Ministry of Health, Rarotonga/Cook Islands; Cook Islands Ministry of Health, Rarotonga Hospital, Rarotonga/Cook Islands; Cook Islands Ministry of Health, Rarotonga Laboratory, Rarotonga/Cook Islands; Cook Islands Ministry of Health, Aitutaki Laboratory, Aitutaki/Cook Islands; Northern Provincial Hospital Laboratory, Espiritu Santo/Vanuatu; Department of Medical Statistics, RWTH Aachen Technical University, Aachen/Germany. Pauwelsstraße 19, 52074 Aachen, Germany; Center of Biostatistics and Epidemiology, Medical School, Sigmund Freud University, Vienna, Sigmund Freud Platz 3, 1020 Vienna, Austria; Central Veterinary Research Institute (CVRI), Ministry of Fisheries and Livestock, P.O. Box 33980, Lusaka, Zambia

## Abstract

**Background:** Arthropod-borne diseases pose a significant and increasing risk to global health. Given its rapid dissemination to all inhabited continents, initiating large-scale outbreaks with severe human infections and economic loss, the Chikungunya virus (CHIKV) is one of the most important arboviruses worldwide. Despite its significance, the real global impact of CHIKV remains underestimated as outbreak data are often incomplete and solely based on syndromic surveillance. During 2011-2016, the South Pacific Region was severely affected by several CHIKV-epidemics, still the area is underrepresented in arboviral research.

**Methods:** 465 outpatient serum samples collected between 08/2016 and 04/2017 on three islands of the island states Vanuatu (Espiritu Santo) and the Cook Islands (Rarotonga, Aitutaki) were tested for anti-CHIKV specific antibodies using Enzyme-linked immunosorbent Assays.

**Results:** A total of 30% (Cook Islands) and 8% (Vanuatu) of specimens were found positive for anti-CHIKV specific antibodies with major variations in national and intranational immunity levels. Seroprevalence throughout all age groups was relatively constant. Comparing the different study settings, four potential outbreak-protective factors were identified: presence of Ae. albopictus (in absence of ECSA E1-A226V-mutation CHIKV), as well as low levels of local population densities, residents’ travel activity and tourism.

**Conclusion:** This is the first seroprevalence study focussing on an arboviral disease in the Cook Islands and Vanuatu. It highlights the impact of the 2014/2015 CHIKV epidemic on the Cook Islands population and shows that a notable part of the Vanuatu test population was exposed to CHIKV although no outbreaks were reported. Our findings supplement the knowledge concerning CHIKV epidemics in the South Pacific Region and contribute to a better understanding of virus dissemination, including outbreak modifying factors. They may support preventive and rapid response measures in affected areas, travel-related risk assessment and infection identification in returning travellers.

**Author Summary:** Mosquito-borne viral infections pose an increasing risk to global health. As they mainly circulate in tropical regions comprised of low-resource countries and affect already vulnerable populations, these diseases have an immense impact on the socio-economic sector. Worldwide, the Chikungunya virus (CHIKV) represents one of the most important arboviruses. Yet, despite its significance, reliable data to understand many populations’ real disease burden is still missing. One example is the South Pacific Region (SPR), which has been severely affected by numerous CHIKV outbreaks since 2011. Epidemiological data is indispensable for the implementation of strategies for optimal allocation of limited resources, efficient early intervention, vector control as well as to understand the virus’ geographical distribution and its contribution to global morbidity. We therefore conducted a CHIKV seroprevalence study in the local populations of two Pacific Island states – the Cook Islands and Vanuatu. Our results show that during the 2014/2015 CHIKV epidemic on the Cook Islands about 30% of the local population has been affected. Although no outbreak was ever officially reported in Vanuatu, a notable part of the respective test population was proven to have been exposed to CHIKV as well. By comparing our two study settings we further identified several environmental and social factors with potential outbreak modifying effect.

## 1. Introduction

Arboviral infections are a significant and increasing challenge to global health [1]. Despite their immense effect on public health and society, including economy and social structures, exact information concerning the real local and global disease burden is often missing [2]. Besides the two important arboviruses Dengue Virus (DENV) and Zika Virus (ZIKV), a third entity adds to the challenges many regions’ health systems must face: The Chikungunya Virus (CHIKV) - an enveloped, RNA Alphavirus belonging to the family of Togaviridae [3]. CHIKV appears in the three distinct phylogroups West African (WA), East/Central/South African (ESCA), and Asian [4] and is transmitted through bites of *Aedes* spp. mosquitoes, mainly *Aedes aegypti* and *Aedes albopictus* [5]. Starting in 2005, large-scale outbreaks in the Indian and the Pacific Ocean area as well as on the American continent, not only highlighted the virus’ potential to cause epidemics of hitherto unknown extent, but also revealed severe complications linked to human infection including crippling and persisting arthralgias, neurological disorders and cardiovascular manifestations [6],[7],[8].

One of the areas severely affected by CHIKV, but still largely missing on the epidemiological map, is the South Pacific Region (SPR) [8],[9],[10]. Historically, the area has been free of CHIKV [11], but since its introduction in 2011 the virus disseminated throughout the region causing numerous outbreaks (Fig 1). The SPR represents a very distinct geographical setting: It is characterized by a vast area of open ocean with thousands of islands scattered in between that comprise the 22 Pacific Island Countries and Territories (PICTs) [12]. These can be subdivided into the three subregions Melanesia, Micronesia, and Polynesia and are home to approximately 11.4 million pacific islanders [13]. Many PICTs fall into the United Nations Development Programme’s categories *developing* or *least developed countries* and are ranked among the countries most at risk from natural disasters [12]. The combination of its archipelagic geography, tropical climate, presence of potent vectors, naive populations, development status and high frequent population flows including millions of tourists each year, makes the region not only particularly vulnerable to arboviral epidemics but also provides nearly ideal outbreak conditions[9],[10],[13],[14],[15],[16],[17]. Despite a high local disease burden, the SPR is therefore assumed to have greatly contributed to the global spread of DENV, ZIKV and CHIKV [9],[10] and a wider knowledge about its arboviral situation is of international interest.

**Fig 1:**
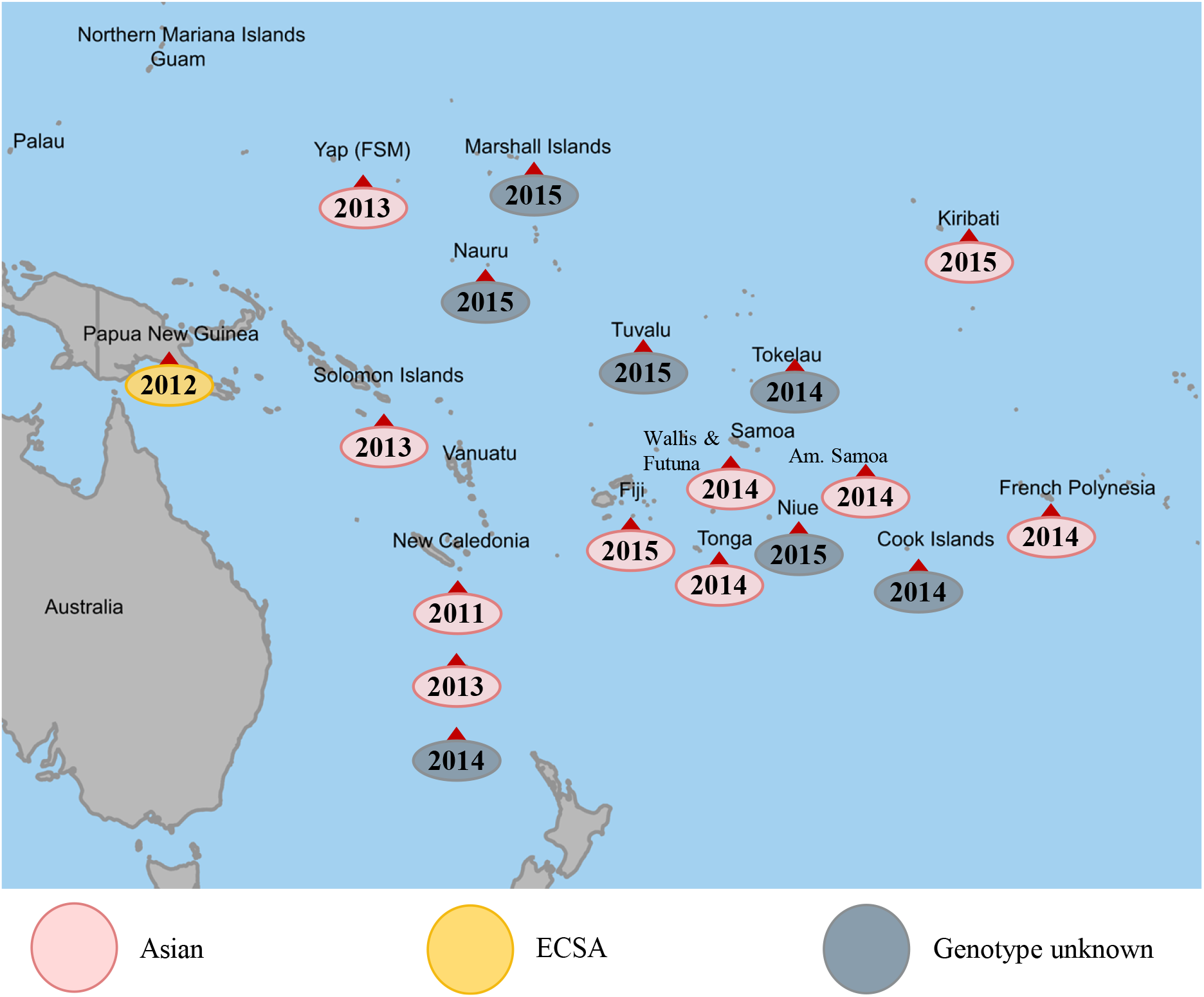
History of CHIKV outbreaks in the SPR, including genotype if known. Source: [10],[13],[17],[18],[19],[20],[21],[22],[23],[24],[25],[26]

We conducted a seroprevalence study in the local populations of two island states: The Cook Islands which were affected by CHIKV in 2014/2015 and Vanuatu that had never reported any CHIKV epidemic. To our knowledge, this is the first seroprevalence study focussing on an arboviral disease in both island states. Further, comparing our two study settings, several outbreak modifying environmental variables were assessed.

## 2. Materials and Methods

Serum samples were collected in hospital laboratories of the island states Vanuatu (outer island Espiritu Santo) during August 2016 – January 2017 and the Cook Islands (main island Rarotonga and outer island Aitutaki) during January 2017 – April 2017. Residents requiring a blood testing within the hospitals’ normal diagnostic routine were asked to participate in the study, excluding tourists and short-term visitors. After written informed consent was given, basic epidemiological information was collected via questionnaires and interviews. 465 samples (197 from Espiritu Santo, 208 from Rarotonga and 60 from Aitutaki) were analysed for anti-CHIKV specific antibodies using a recombinant-antigen-based indirect Enzyme-linked immunosorbent Assay (ELISA) (EUROIMMUN (EI 293a-9601 G); sensitivity: 95.4%; specificity: 98.6%). Test results were defined as “positive”, “negative” or “equivocal” if in-between the threshold values. The study was conducted in accordance with the Ethics Committee of the medical faculty of the RWTH Aachen University (agreement 051/16_09/05/2016) and with the local authorities of the Cook Islands (Ref.: #16-16) and Vanuatu (Ref.: MOH/DG 10/1/1-GKT/lr).

Associations between ELISA test results and gender as well as between seroprevalence levels and test collective were performed by means of Chi^2^-Test. To test for associations between ELISA test results and age (age groups ‘adults’ (>15 years) and ‘children’ (0-15 years)), the Fisher-Freeman-Halton Exact Test was used due to smaller test collectives and percentages of >20% of expected frequencies below 5. For all comparisons the significance level was set to 5%; due to the explorative nature of the investigation no adjustment to the significance level was made. Results were reported as percentage and two-sided p-values. For estimation and comparison of community immunity-levels, raw data were directly standardized by age (10-year age groups; excluding the age group of 0-9 years due to low numbers) and gender according to the standard populations “total resident population” (Cook Islands) and “total population living in private households” (Vanuatu) as published in the Cook Islands Census of Population and Dwellings, 2011 [27] and the Vanuatu Post-Tropical Cyclone Pam Mini-Census, 2016 [28] respectively (further referred to as “resident population” or “residents”). Estimates of the immunity in the different collectives were accompanied by 95% confidence limits (Cl) and p-values.

Information concerning potentially outbreak-modifying environmental and social factors was obtained from local authorities as well as from the DWD Climate Data Center. Data regarding the Cook Islands CHIKV outbreak derive from the archives of the Pacific Public Health Surveillance Network. All analyses were performed with Microsoft Excel Office 365 and IBM SPSS Statistics 21.

## 3. Results

Important epidemiological, environmental, and social data of the different study sites are displayed in table 1.

**Table 1:**
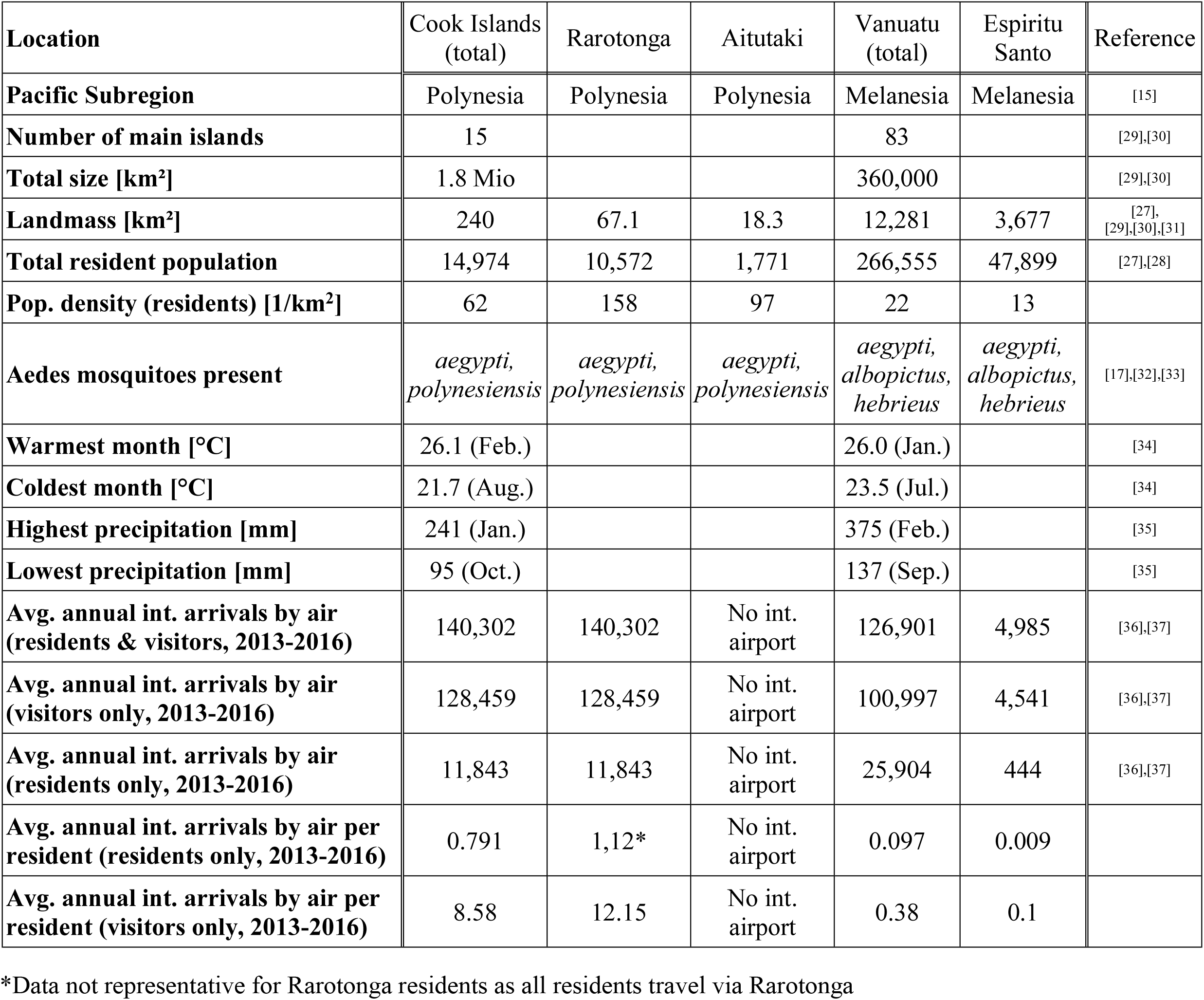
Environmental and social data of epidemiological relevance in the study sites

According to the origin of the specimen, we mainly considered 2 different test populations in our analysis: The Cook Islands and the Vanuatu collective. The Cook Islands collective can be further divided into two subgroups corresponding to the different islands of origin, Rarotonga and Aitutaki (Table 2).

**Table 2:** Test population structure – gender distribution and median age

| **Location** | **Both sexes**<br>**[No**<br>**samples]** | **Male**<br>**[No**<br>**samples]** | **Male**<br>**[%]** | **Female**<br>**[No**<br>**samples]** | **Female [%]** | **Median age**<br>**[years]** |
| --- | --- | --- | --- | --- | --- | --- |
| Cook Islands | 268 | 122 | 45.5 | 146 | 54.5 | 49 |
| Rarotonga | 208 | 83 | 39.9 | 125 | 60.1 | 47 |
| Aitutaki | 60 | 39 | 65 | 21 | 35 | 53,5 |
| Vanuatu (Espiritu Santo) | 197 | 80 | 40.6 | 117 | 59.4 | 36 |
| Total | 465 | 202 | 43.4 | 263 | 56.6 | --- |

Anti-CHIKV specific antibodies could be found in 30.2% (81 individuals) of all Cook Islands specimen and in 8.1% (16 individuals) of the Vanuatu test collective. Further subdividing the Cook Islands test group, a larger percentage of the Rarotonga specimen was identified as positive (33.2%; 69 individuals), compared to the serum samples from Aitutaki (20%, 12 individuals) (Fig 2).

**Fig 2:**
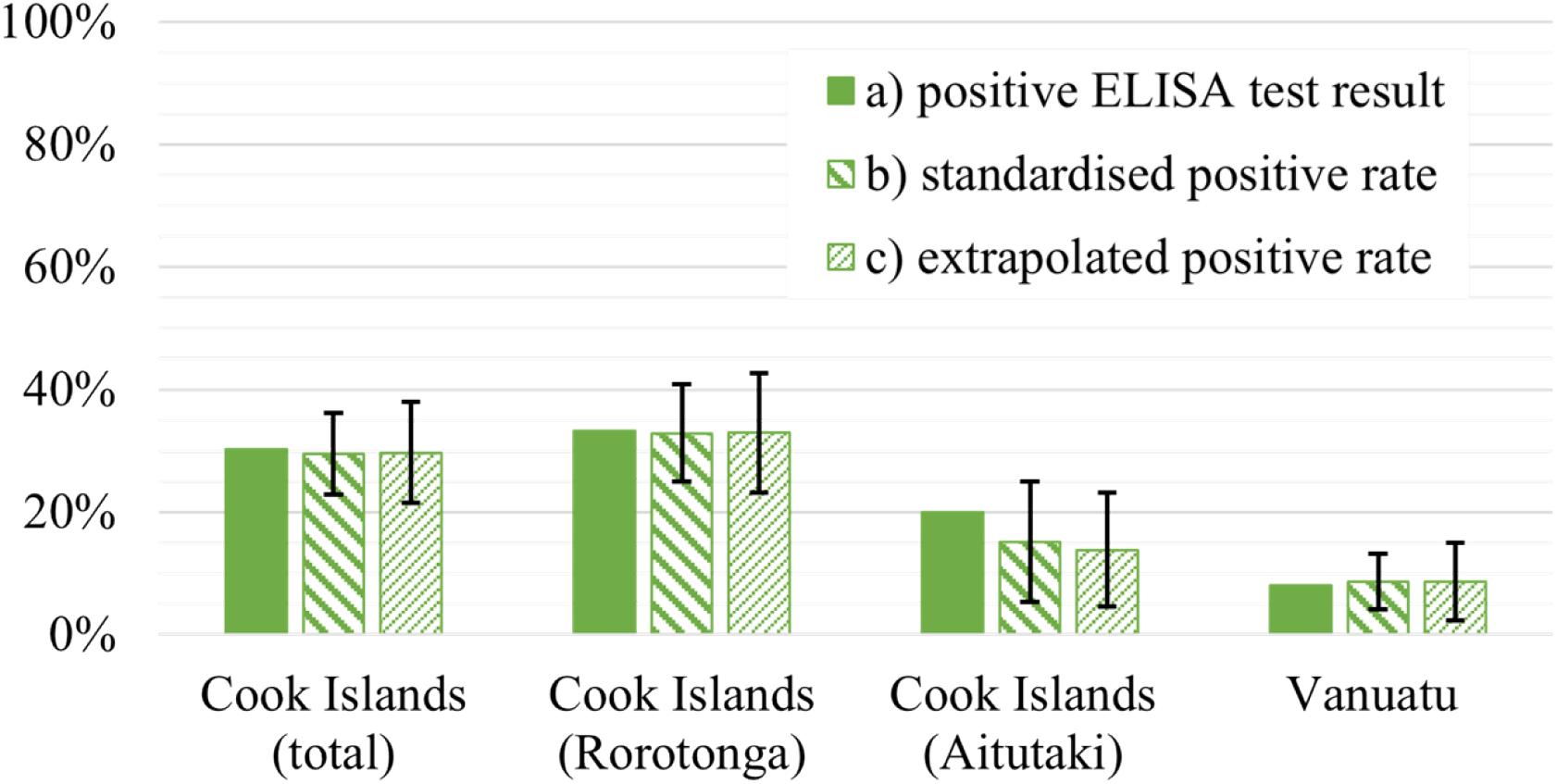
a) positive ELISA test results. b) seroprevalence rates standardized by gender and age. c) seroprevalence rates extrapolated to the total resident population. Black whiskers show 95%CI relative to standardized and extrapolated rate.

Using the Chi^2^- and the Fisher-Freeman-Halton Exact Test respectively, we could show that in our two main test collectives, there was neither a significant association of ELISA test results and gender (Chi^2^-Test, p>0.05) nor significant differences between the age groups ‘adults’ (>15 years) and ‘children’ (0-15 years) (Fisher-Freeman-Halton Exact Test, p>0.05). Instead, seroprevalence levels were relatively constant throughout age groups without variations of statistical significance (Fisher-Freeman-Halton Exact Test, p>0.05) (Fig 3).

**Fig 3:**
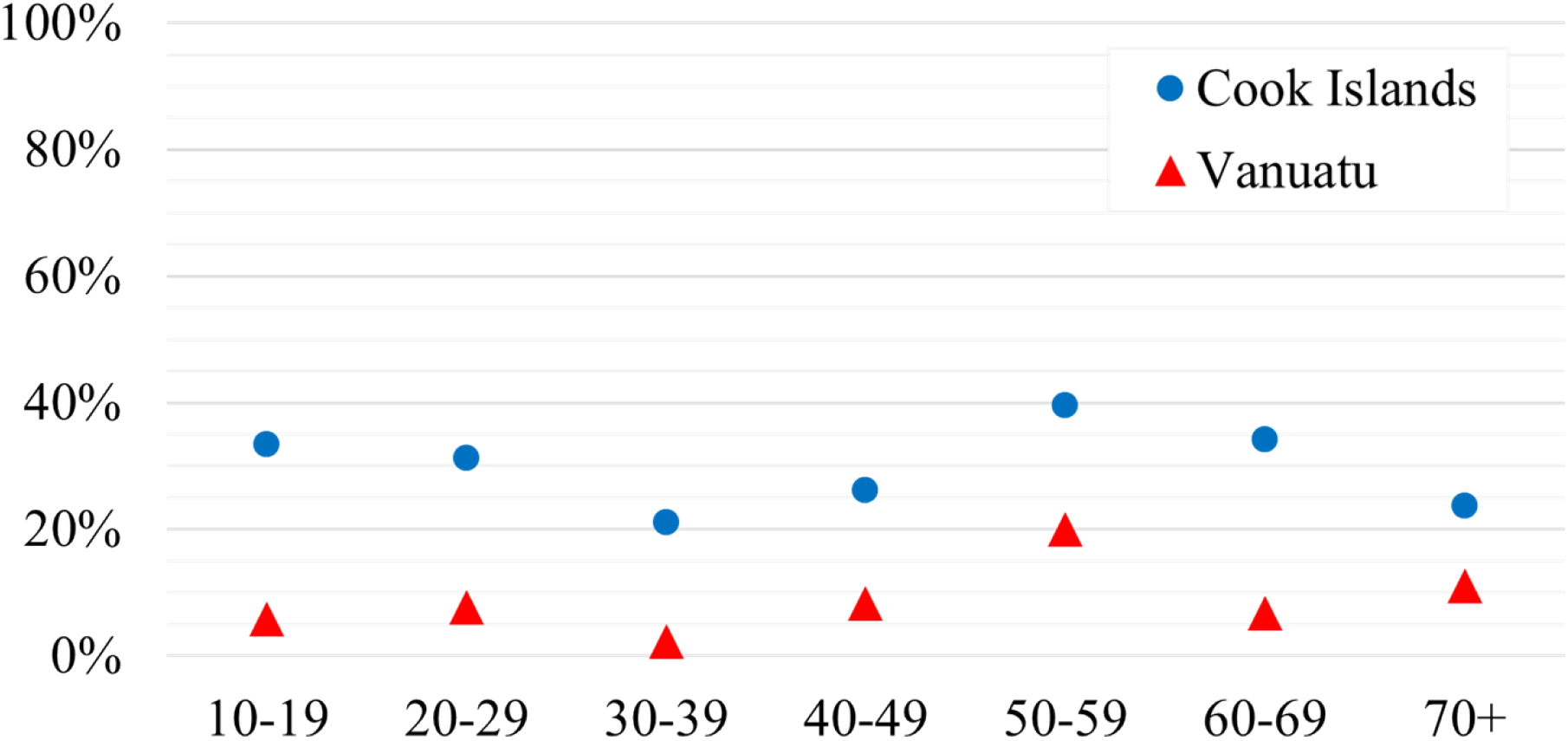
positive ELISA test results by 10 years age groups (excluding age group 0-9 years)

To allow a direct comparison between our test collectives, data were standardized by age and gender (Fig 2) with significant differences in immunity rates for CHIKV between the Cook Islands (29.5%; 95%CI: 22.9-36.1) and the Vanuatu test collectives (8.6%; 95%CI: 4.1-13.2) (Chi^2^-Test, p<0.05). Differences in seroprevalence levels were also significant between our two Cook Islands sub-populations from Rarotonga (32.9%; 95%CI: 24.9-40.8) and Aitutaki (15.1%; 95%CI: 5.3-25) (Chi^2^-Test, p<0.05). Extrapolated to the total population of both island states, calculated immunity levels sum up to 29.7% (95%CI: 21.4-38) in the Cook Islands’ total population and to 8.6% (95%CI: 2.2-15) in Vanuatu with significant difference (Chi^2^-Test, p<0.05) (Fig 2). In the two Cook Islands sub-populations, extrapolated rates amount to 32.9% (Rarotonga) (95%CI: 23.2-42.6) and 13.8% (Aitutaki) (95%CI: 4.5-23.1) as well showing a significant difference (Chi^2^-Test, p<0.05).

The Chikungunya epidemiological history of the Cook Islands epidemic is displayed in Fig 4. Between 10/2014 and 08/2015 there were 18 confirmed and 782 clinically diagnosed cases [13],[38]. Considering our extrapolated results this corresponds to a calculated case-detection-rate of 18% for this specific outbreak.

**Fig 4:**
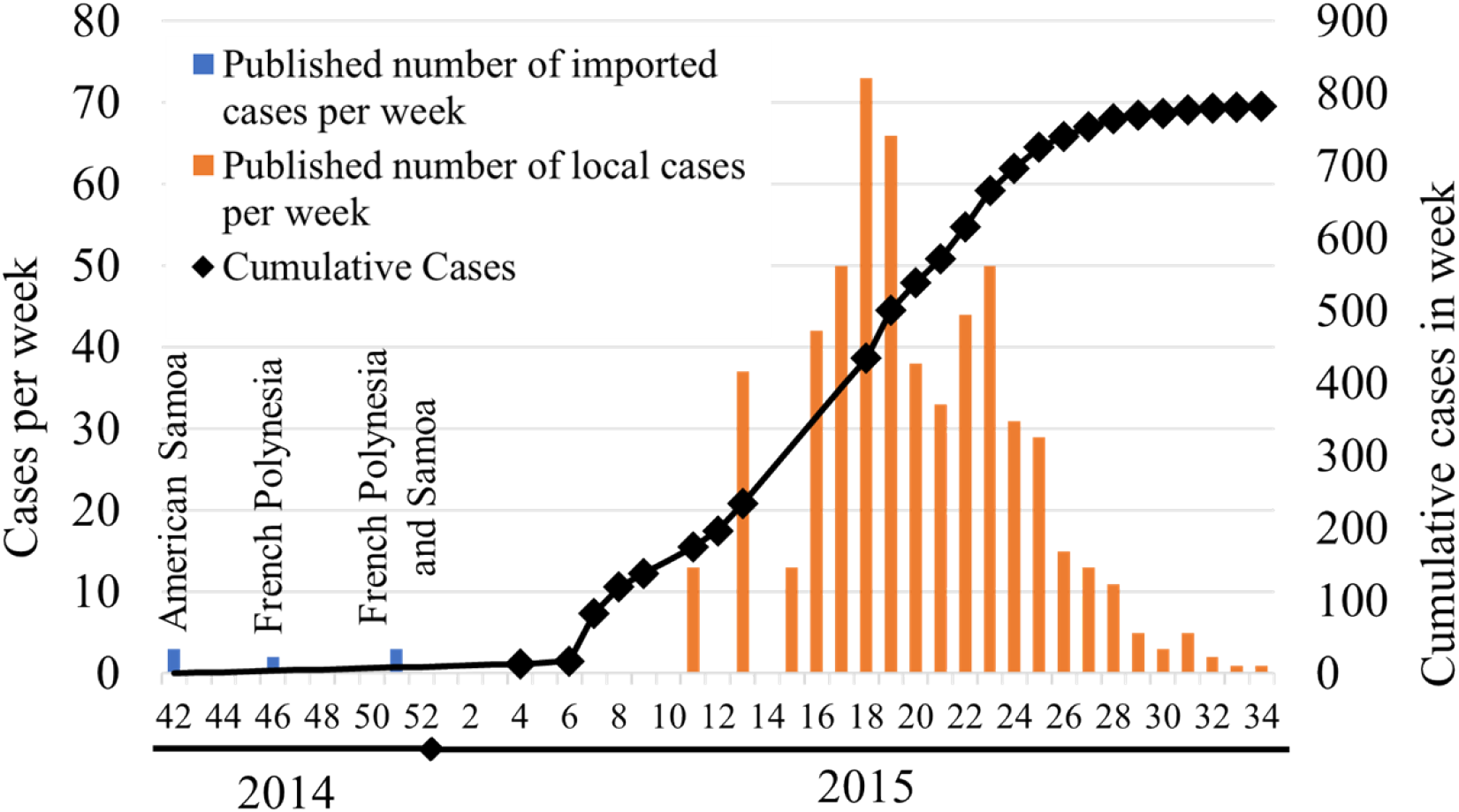
2014/2015 Cook Islands CHIKV outbreak as reported by local syndromic surveillance systems. Source: [26]

## 4. Discussion

Seroprevalence surveys are an indispensable tool for exact estimations of arboviruses’ health impact, geographical distribution and trends in transmission [2]. Investigating the effects of the 2014/2015 Cook Islands CHIKV epidemic, 30% of our test population showed evidence for prior CHIKV infection. Significant differences between both subgroups (Rarotonga and Aitutaki) depict the effects of a scattered archipelagic geography on arboviral dissemination and are in line with surveillance data: During the outbreak hundreds of clinical cases were reported on the main island Rarotonga, but only five other islands, including Aitutaki, registered some infections and except for one, all index cases had travel history to Rarotonga [39]. The absence of significant differences between seroprevalence rates in children and adults as well as relatively little variation in immunity rates among 10-years age groups, suggest the primal epidemic and self-limiting occurrence of CHIKV in the Cook Islands, again consistent with surveillance data[26]. However, surveillance data are especially useful for outbreak detection but fail to depict the real disease burden in a population [2]. They often remain incomplete, sporadic or delayed and are known to underestimate the dimensions of an epidemic [2],[13] - an effect once again demonstrated by our calculated case-detection-rate of 18% for the Cook Islands CHIKV outbreak.

Although Vanuatu never reported CHIKV circulation, our data provide evidence that 8% of the test collective were previously exposed to the virus. Seropositivity was most likely acquired during the phase of multiple outbreaks in the SPR, and infections may have occurred either abroad and travel-related or during very localized outbreaks that never reached epidemic extent. Explanatory approaches for the differences in seroprevalence levels observed in this study require the investigation of environmental and social driving factors of arboviral spread. For this purpose, we compared our study geographical settings in terms of vectors, climate, population density, mobility of the residential population and tourism (table 1). Both settings host the competent vector *Ae. aegypti*. In terms of secondary vectors, the Cook Islands are home to *Ae. polynesiensis* while in Vanuatu *Ae. albopictus* can be found [17],[32]. Another species of the genus *Aedes* present in Vanuatu is *Ae. hebrideus* [32], but as for this mosquito data concerning CHIKV transmission are scarce it is not further discussed within this work. Regarding the epidemiological effect of the two secondary vectors *Ae. albopictus* and *Ae. polynesiensis*, there are, however, some major differences: *Ae. polynesiensis* is particularly well-suited to PICT ecosystems as it uses artificial as well as a variety of natural breeding grounds including highly specific biotopes such as land crab burrows and fallen coconuts [32]. It therefore occurs in multiple environments and is very likely to act synergistically to *Ae. aegypti* in virus dissemination [32],[40],[41]. In contrast, *Ae. albopictus* has occasionally been shown to replace the main vector *Ae. aegypti* [42]. Further, CHIKV transmission by *Ae. albopictus* mosquitoes is directly associated with an E1-A226V mutation in the ECSA-CHIKV envelope protein [43], while the Asian CHIKV lineage in general is limited in its ability to adapt to this vector [44]. Thus, *Ae. albopictus*’ status as secondary vector in Vanuatu is CHIKV genotype-dependent, and due to its tendency to replace *Ae. aegypti*, high numbers of *Ae. albopictus* could even be regarded as a factor supporting a reduced and weak transmission cycle, provided that there is no introduction of the ECSA E1-A226V-mutation CHIKV.

In terms of climate, both the Cook Islands and Vanuatu offer favorable conditions for vectors: Throughout the year maximum and minimum of monthly means of air temperature as well as monthly means of precipitation are close to the hypothesized CHIKV transmissibility optimum of 25°C and 206mm respectively [45] (table 1). We therefore conclude that neither temperature differences nor varying precipitation can be identified as crucial factors to the disparate seroprevalence levels. Population densities on the other hand have shown to be concordant to detected seroprevalence levels (table1).

As CHIKV has demonstrated its potential to spread via airline routes [21],[46],[47],[48],[49],[50] we further examined differences in the two island states’ migration and tourism profiles: In general, Pacific islands communities share close social and economic connections, both within individual countries and international including high bidirectional population flows and millions of tourists visiting each year [12],[15]. There are however distinct variations in tourism and country specific mobility patterns between the different subregions: Polynesian islands show strong individual mobility and high international migration rates while in many Melanesian and Micronesian Island states, population movements arise predominantly within the country and international migration is at a low level [15]. Data concerning international arrivals by air (the main travel route) highlight the much higher number of individual international travels and arriving tourists per resident on the Cook Islands compared to Vanuatu (table 1). Considering this, the Cook Islands CHIKV outbreak in 2014/2015, following a series of epidemics in the region, is not surprising and it can be assumed that the virus’ spread to the outer island Aitutaki was facilitated by intensive (touristic) travel activities to and from the main island Rarotonga [51]. On the other hand, lower rates of residents’ mobility and tourism in Vanuatu probably contributed to the limited seropositivity detected in our survey despite an estimated high risk of virus importation [21].

Summarizing, this is the first seroprevalence study focusing on an arboviral disease in the Cook Islands and Vanuatu. It highlights the impact of the 2014/2015 CHIKV epidemic on the Cook Islands population and shows that a notable part of the Vanuatu test population had previous CHIKV exposure although no outbreak was reported before this study. Comparing the different study settings, our results underline the effect of environmental and social factors on CHIKV dissemination in Pacific Islands settings and four factors potentially reducing Vanuatu’s epidemic risk within the scope of CHIKV dissemination throughout the SPR were identified. These factors include presence of invasive Ae. albopictus (if no ECSA E1-A226V-mutation CHIKV is introduced), low population densities, low travel activity of the local population, and moderate tourism.

Our findings supplement and correct the knowledge concerning CHIKV epidemics in the SPR which is often based on incomplete surveillance data. In addition, our findings may support preventive and rapid response measures in affected areas and are of international interest for travel-related risk assessment and infection identification in the era of globalization.

As with many seroprevalence surveys, limitations result from the study design [2]. Representativeness is lowered by using serum samples collected from hospital patients, rather than from the general population. Further, not all islands of the two island states could be included in the analysis. Regarding the isolated character of our study settings, this could lead to wrong estimations concerning the seroprevalence levels on islands not depicted in this survey and extrapolated seropositivity rates should be interpreted with caution.

Another limiting factor which cannot be ruled out is the risk of cross-reactions of antibodies against other alphaviruses, e.g., the Ross River Virus (RRV). RRV emerged in the SPR in 1979-1980, causing large outbreaks with evidence for silent circulation in several PICTs ever since [52],[53]. However, this hypothesis seems rather unlikely: In addition to the fact that there was no known RRV epidemic in Vanuatu in the past, the remaining seroprevalence levels would be expected to be higher in age groups already born during the epidemic (40+ years) and to rise over time in age-dependent manner in case of silent circulation.

## Data Availability

See file "supplemental data"

## Acknowledgements

Our special thanks go to Dominik Mildt for technical editing and continuous support. We further thank DHL Cook Islands and DHL Vanuatu for logistical assistance.

## Supporting information captions

S1 Test population and ELISA test results_CHIKV

## Notes

### Competing Interest Statement

The authors have declared no competing interest.

### Funding Statement

The study was supported by: Aachen Dental and Medical Expeditions (ADEMED e.V. https://www.ademed.de/), a non-profit society to support research in travel medicine [grant number 180627] (Author: CS, funding: financial). The funders had no role in study design, data collection and analysis, decision to publish, or preparation of the manuscript. and by the German Army Institute of Microbiology (Author: CS, funding: materials and use of their laboratory [no grant / grant number]). The funders had no role in study design, data collection and analysis, decision to publish, or preparation of the manuscript.)

### Author Declarations

Study was approved by the ethical committees of RWTH Aachen University (051/16_09/05/2016) and with the local authorities of the Cook Islands (Research permit reference number #16-16) and Vanuatu (Ref.: MOH/DG 10/1/1-GKT/lr).

